# Assessing the Impact of Extreme Weather Events on HIV and Tuberculosis Care in Zimbabwe

**DOI:** 10.64898/2025.12.22.25342867

**Authors:** Canice Christian, Rutendo Mukondwa, Cesar Aviles-Guaman, Mollie Hudson, Kudakwashe Takarinda, Nora West, Amrita Ayer, Karen Webb, Priya B. Shete

## Abstract

The growing threat to health systems posed by extreme weather events (EWEs) disproportionately affects people living with HIV (PLHIV) and tuberculosis (TB) in high-burden, low-resource settings. In Zimbabwe, for example, the 2023/2024 El Niño-induced drought intensified food insecurity and reduced access to clinics and medication for vulnerable populations. We conducted a formative evaluation to assess how recent EWEs in Zimbabwe have impacted HIV and TB care in this community and to identify potential climate-adaptive health-service strategies. First, we surveyed 1,804 PLHIV across 321 clinics in Zimbabwe to describe their perceptions of EWEs on their health. We conducted focus group discussions (n=6) with HIV and TB affected individuals. The majority of survey participants (76%, 1363/1804) reported noticing changes in climate over the past 5 years with 75% (n=1024) reporting disruptions to their HIV care due to these changes. Survey respondents reported climate-related disruptions led to increased food insecurity (86%, 877/1024) and water shortages (65%, 667/1024) that were dominant barriers to treatment adherence. Qualitative analysis confirmed barriers to HIV and TB care from extreme weather events included widespread food insecurity, crop failures and livestock deaths, with consequent impacts on income leading to challenges paying for food and transportation to clinics. Participants highlighted the need for expanded interventions that facilitate social and economic supports, including nutrition support, water access, transportation, and income-generating activities, as well as decentralized models of healthcare delivery. Findings underscore the urgent need for integrated, climate-adaptive strategies to maintain HIV and TB care continuity in climate-vulnerable regions like Zimbabwe.

**Panel: Research in Context:** **Evidence before this study**
Previous studies have shown that climate shocks such as droughts, flooding, and heat waves have important consequences for social and structural determinants of health, including food insecurity, water access, livelihoods, and mobility, and that these determinants can influence HIV and TB risk and outcomes. However, almost all available evidence has been conceptual rather than empirical. Few studies have examined patient-reported impacts of extreme weather events on HIV or TB care, and none have assessed climate-adaptive health-service strategies identified directly by affected communities and frontline health workers in Zimbabwe or similar high-burden, climate-vulnerable settings.
**Added value of this study**
This study provides formative empirical evidence on how extreme weather events disrupt HIV and TB care from the perspective of people living with HIV, people affected by TB, and health-care workers. Using a large multi-district patient survey alongside qualitative data, we document how climate-related food insecurity, water scarcity, financial strain, mental health stress, and transportation barriers undermine treatment adherence and continuity of care. Further, the study identifies community-prioritized strategies to sustain HIV and TB care during climate shocks, including nutrition and water support, transportation assistance, decentralized service delivery, and livelihood interventions.
**Implications of all available evidence**
As climate change intensifies, strategies that protect continuity of HIV and TB care will be increasingly essential in high-burden settings. Findings suggest that social protection and decentralized health-service models may mitigate climate-related disruptions to treatment, promote resilience, and reduce the structural burden placed on patients. Integrating climate-adaptive approaches into HIV and TB programming could strengthen health-system preparedness and promote sustainable long-term outcomes for climate-vulnerable populations.

## Introduction

Climate change-associated extreme weather events (EWEs) increasingly threaten the health and wellbeing of people living with HIV (PLHIV) and tuberculosis (TB), particularly in high-burden, resource-limited settings. EWEs, such as extreme heat, drought, and excessive rainfall, amplify social and structural determinants that heighten susceptibility to HIV and TB and worsen clinical outcomes among populations already facing multiple vulnerabilities.^1^ Several conceptual frameworks describe how climate-sensitive pathways such as food insecurity, undernutrition, health system disruption, and population displacement influence TB transmission, TB disease progression, and HIV acquisition risk.^1,2^ For example, Lieber *et al* (2021) proposed a model linking climate change and HIV outcomes, identifying food insecurity as a key mediator of increased HIV risk in the context of climate-related shocks.^2^ Similar mechanisms apply to TB, where climate-related food insecurity compromises nutrition and treatment outcomes.^3,4^ Other fundamental determinants of TB including poverty, inadequate access to health services, forced migration, and occupational exposures, are also influenced by EWEs, although the magnitude and pathways of these associations remain insufficiently characterized.^5^

Building on these conceptual models, there is an urgent need for empirical evidence that quantifies the impact of EWEs on HIV and TB programs. Such data include the influence of changing social, environmental, and structural conditions on service delivery, program implementation, and patient outcomes.^1^ Despite growing concern, few studies have systematically examined how climate hazards affect the operational performance of HIV and TB programs or the lived experiences of affected communities.

Zimbabwe represents a compelling case study of the convergence of climate vulnerability and dual HIV/TB epidemics. HIV/AIDS and TB remain leading causes of mortality in the country^6^ despite significant progress made in ending the HIV epidemic through surpassing the 95-95-95 goals^7^ and reducing TB incidence nationally^8^. However, recent EWEs, including prolonged droughts, excessive rainfall, and episodic flooding, have exacerbated existing social and health system pressures and may reverse progress in the HIV and TB epidemics. The 2023-2024 El Niño-induced drought led to severe reductions in agricultural production, driving widespread food insecurity.^9,10^ Projections suggest that climate-related disruptions in Zimbabwe will intensify in the years ahead, further imperiling already fragile health and social systems.^11^

The goal of this study was to explore the perceived impact of climate change and EWEs on HIV and TB determinants, service delivery, and clinical outcomes, as reported by affected communities, in addition to identifying community-prioritized mitigation strategies with the potential to feasibly address climate-sensitive determinants and barriers to care.

## Methods

We conducted a survey-based formative evaluation supplemented by qualitative data. We conducted surveys with a sample of PLHIV and people affected by TB. We also conducted focus group discussions (FGDs) with people affected by TB and PLHIV among a sub-sample of those surveyed to gain further insights into the impacts of EWEs on HIV and TB service delivery and to identify implementation strategies to bolster climate adaptation and resilience.

### Study setting

The Organization for Public Health Interventions and Development (OPHID) is a health service delivery organization specializing in HIV and associated care. Through collaboration with public health and community partners in Zimbabwe, OPHID aims to enhance access for communities to comprehensive HIV and TB prevention, care and treatment. Study activities were conducted within OPHID’s network of >600 health facilities in Zimbabwe. These facilities range from rural, urban, district, provincial, mission to polyclinics and central hospitals providing comprehensive HIV and TB prevention, care and treatment services within the public health system.

### Surveys

We collected survey and clinical data among people living with HIV (PLHIV) at a sample of clinics in Zimbabwe. We surveyed PLHIV (> 18 years) on antiretroviral therapy (ART) from November–December 2024 using Client Satisfaction Surveys across 321 OPHID-supported health facilities in 15 districts in Zimbabwe. These surveys are a standard practice used by OPHID to identify issues affecting the care. The PLHIV surveyed were selected using random sampling. The OPHID District Community Officer and the Zimbabwe National Network of People Living with HIV District Coordinator identified eligible clients who had accessed opportunistic infection or ART services that month and randomly selected participants for interview by trained Community HIV and AIDS Support Agents (CHASA). Exclusion criteria included not receiving HIV care at an OPHID clinic or age <18 years.

Surveys included questions related to the impacts of EWEs in their communities, how EWEs have impacted HIV/TB care, what, if any, services are available to mitigate climate impacts on health, and what people affected by HIV and TB believe are strategies that could better address challenges related to service delivery given the impacts of climate change. The survey instrument was created in conjunction with local healthcare worker staff and researchers, drawing elements from validated instruments and adapted to the local context. The instrument was piloted to assess clarity, relevance, and flow and refinements were made based on feedback.

### Focus Group Discussions

We conducted a total of six focus group discussions (FGD) across three districts in Zimbabwe (Bulilima, Mangwe, Matobo) with a range of 5-8 participants per FGD. Sample size was based on a systematic review of data saturation in qualitative studies that found that in as few as 4 FGDs reached saturation.^12^ In each district, we conducted one FGD among people affected by TB (defined as people actively or previously received TB treatment at the facility) and one FGD among PLHIV. We employed a convenience sampling approach to select patients who had visited the health facility for medical check-ups, medication resupply, or viral load testing, as recorded in clinical registers and verified by clinical staff. The interviews were conducted in Ndebele or Shona and one study staff recorded summary memos and key quotations. Interview guides can be found in Appendix A.

### Analysis

Quantitative data were analyzed using descriptive and summary statistics with Stata V16.1.^13^ For the qualitative analysis, memos and key quotes were summarized following each FGD by a trained qualitative researcher based at OPHID (RM). The findings from the FGDs with those affected by TB and PLHIV were compared when possible to elicit differences between the two groups. General themes from the qualitative data were summarized using a modified framework approach^14^ and mapped to survey results, when possible.

Themes were then mapped to the Consolidated Framework for Implementation Research (CFIR). This implementation science framework describes the determinants of implementation efforts across domains of the outer setting, inner setting, and individual characteristics.^15^ We used a local needs assessment strategy to identify strategies for mitigating the impacts of EWEs on HIV and TB care. Through FGDs with PLHIV and those affected by TB, participants themselves identified the challenges they face during EWEs and proposed practical strategies to address them. Insights from these discussions helped to identify locally relevant and practical solutions. During qualitative interviews, we systematically asked participants which strategies would be most beneficial for their access to care given challenges related to EWEs.

### Ethics

The study was approved by the Institutional Review Boards of the Medical Research Council of Zimbabwe and the University of California, San Francisco. For the surveys, written informed consent was waived because the surveys were routinely collected during care received at the clinic and the primary purpose was to evaluate satisfaction of care. We obtained written, informed consent from FGD participants. Surveys and transcription of audio recordings were de-identified. FGD participants were compensated $5 USD for their time.

### Statement on Positionality

The team collecting data and conducting the analysis for this paper consisted of researchers from Zimbabwe and the United States who have expertise in HIV and TB care, health systems research, qualitative and quantitative methods. The researcher who conducted the FGDs was a trained social science researcher from Zimbabwe who was aware of the contextual factors of the study population. The surveys were conducted by trained CHASA familiar with the context in which those receiving care at OPHID clinics are living. The data analysis and writing of this manuscript was conducted among a group of skilled researchers and reflexivity involving reflexive memos and team discussions was engaged throughout the data analysis process to enable validation of data interpretation and to engage in understanding how positionality may have influenced our findings.

## Results

A total of 1804 eligible participants completed the survey which represents the total number of individuals who were approached to participate in the 15 districts where the questionnaire was administered (Table 1). No participants were excluded after enrollment and all those who completed the questionnaire were included in the analysis.

**Table 1.**
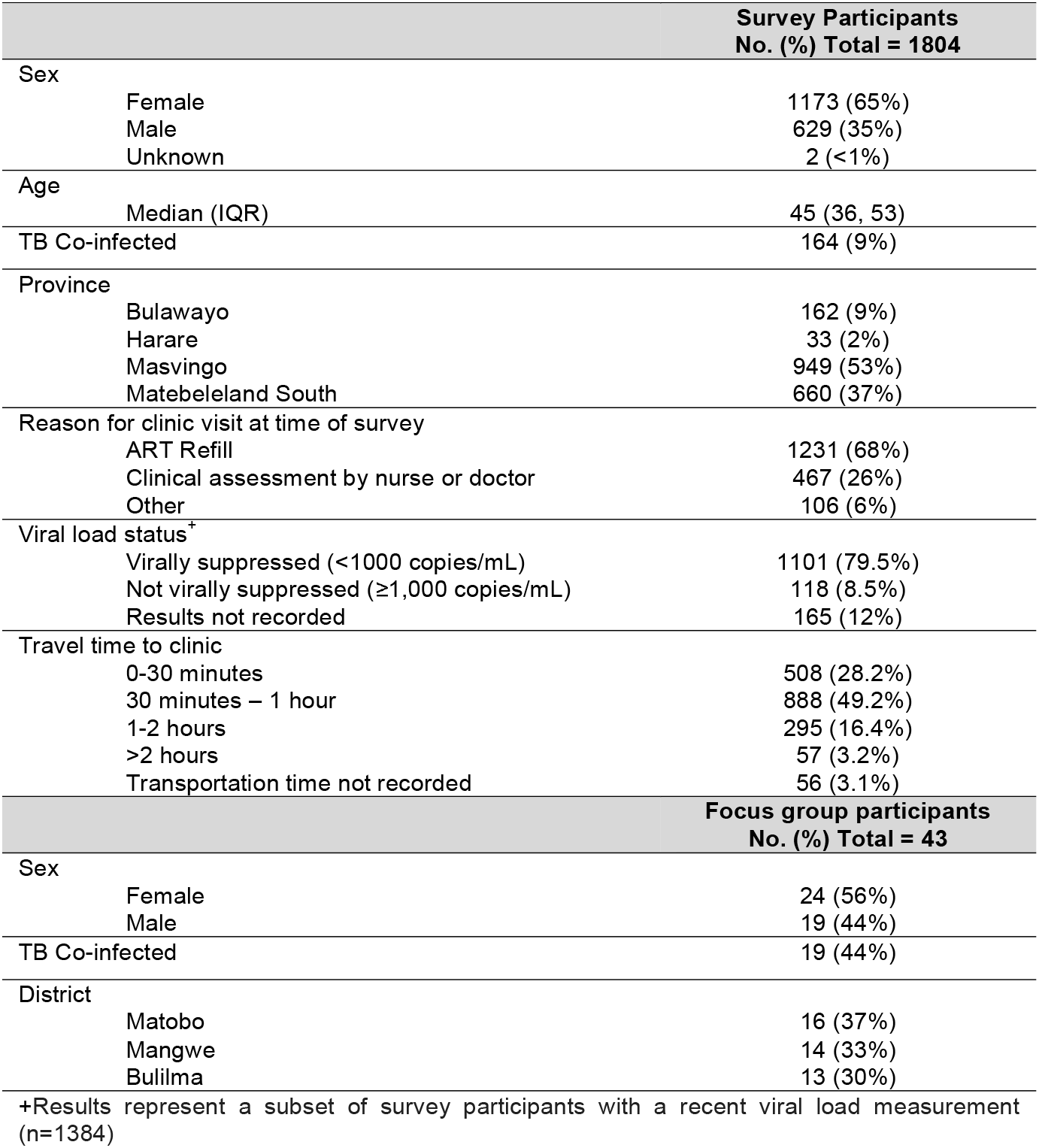
Study participants completing surveys and focus groups evaluating the effects of EWEs on HIV and TB service delivery in Zimbabwe.

### Effects of extreme weather events on TB/HIV care

1363 of 1804 (76%) survey participants reported noticing changes in the typical climate of their communities over the past 5 years. Of those who experienced these changes, 75% reported disruptions to their HIV care due to EWEs/climate change (n=1024). Quotations from qualitative data provide further illustration of how climate and EWEs such as extreme heat impact disruptions to care (Figure 1).

**Figure 1.**
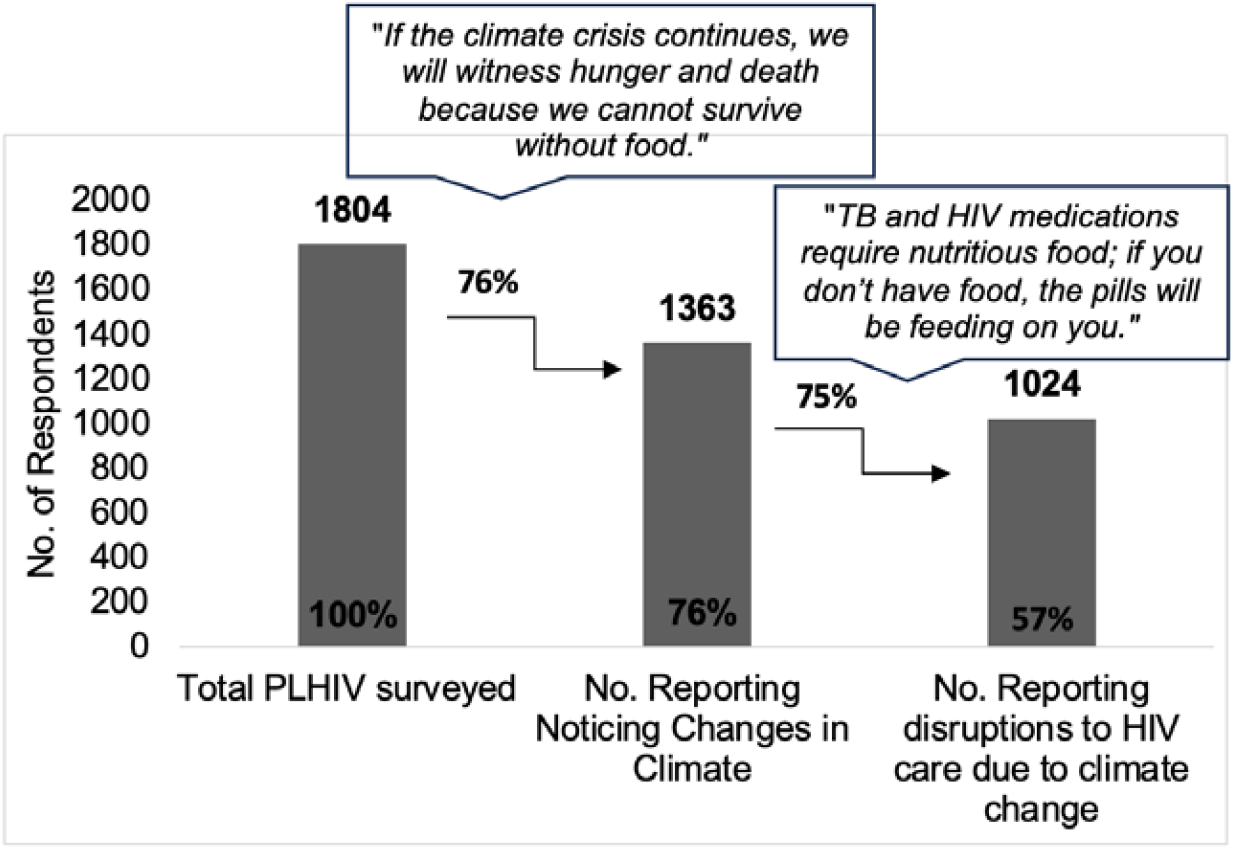
Cascade of the impact of climate change on HIV care among PLHIV receiving care at OPHID clinics in Zimbabwe including the quotes from those impacted by climate change

### Mechanisms of Extreme Weather Event Disruption on TB/HIV Care

Results from surveys and FGDs analyzed using CFIR revealed potential mechanisms by which EWE disruptions impacted TB/HIV care in this population at the individual-, facility and community-(inner-setting), and systems-level (outer setting) levels.

#### Individual-Level Challenges

Individuals effected by HIV and TB reported climate change impacting their mental health (308/1024, 30%) (Table 2). When asked about the impacts of climate change on their HIV and TB care in FGDs, TB patients highlighted the significant impacts of climate change and TB on their well-being and health. Those affected by TB expressed that climate change has introduced a constant source of stress and has exacerbated issues such as fatigue, respiratory problems, food insecurity, and water shortages. Many participants shared the emotional and physical toll, with some experiencing weight loss, malnutrition, and suicidal thoughts due to the ongoing hardships.

**Table 2.** Mechanisms by which climate change and extreme weather events disrupted HIV care for study participants in Zimbabwe (n=1024)

| Unmet needs/vulnerabilities described by participants as resulting from climate change and EWEs: | Study Participants, No. (%) |
| --- | --- |
| Lack of food for you or your family | 877 (86%) |
| Lack of water for you or your family | 667 (65%) |
| Lack of funds for user fees, transport, or other healthcare costs | 468 (46%) |
| Lack of community or social support due to additional stress in the community | 329 (32%) |
| Mental health related issues | 308 (30%) |
| Limited transportation options or transport challenges | 294 (29%) |
| Inability to safely store medications due to extreme temperatures | 287 (28%) |

> *“Being HIV positive it’s a burden on its own, climate change is worsening the burden through stress and it’s putting our life in danger.”* Mangwe FGD PLHIV

Participants also discussed fatigue caused by the extreme heat, which one participant described as *“iyavuthisa umzimba,”* (FGD people affected by TB, Matobo) meaning it feels as though the body is being “cooked” in such extreme temperatures. Many focus group members reflected on consequent difficulties maintaining a consistent routine in these circumstances, resulting in missed doses or improper timing of medication.

#### Facility and Community-Level Challenges

Survey respondents reported EWEs/climate change increasing food insecurity (887/1024, 86%) (Table 2), leading to challenges in treatment adherence and competing priorities. Many respondents mentioned taking their HIV medications without food, leading to negative health consequences such as headaches and fatigue, which contributed to skipping their medication and ultimately worsened adherence.

> *“If you take medication without food, you become so weak that you might not even be able to wake up, and in some cases, you could end up being hospitalized.”* FGD, people affected by TB, Bulilima district.

Participants discussed that changing weather conditions have caused the disappearance of caterpillars, locally known as *‘amacimbi*,’ which are a source of income and protein in the community. Crop failures and livestock deaths, which participants relied on for food and income for transportation to the health facilities, were noted as severe disruptions.

> *“Now, we eat only once a day, like dogs.”* FGD, PLHIV, Bulilima district

Sixty-five percent (n=667) of survey participants cited insufficient water access as a challenge of climate change on their HIV care. During FGDs, participants expressed concern about dwindling water resources, which have forced them to rely on unprotected wells that are unsafe and can lead to diseases like diarrhea and cholera.

“*Climate change has led to water scarcity and we sometimes go up to 3 days without water in the taps and the boreholes are not functional*.*”* FGD, PLHIV, Mangwe District

Transportation and distance to health facilities were an added challenge of climate change on HIV and TB care. Participants reported long distances to healthcare facilities (up to 20 km) during extreme heat complicated their ability to access treatment. In FGDs, participants also described the challenge of crossing rivers during heavy rains, which can delay access to treatment due to flooding.

Forty-six percent of participants identified financial barriers, including the costs of transportation, as critical issues affecting their ability to access care (n=468). During the FGDs, participants reported that the loss of livelihoods due to climate change has made it harder for them to meet basic needs, especially as farming has become unreliable, leading many to rely on labor-intensive, low-paying jobs. Participants also discussed psychological stress resulting from an inability to work and the stigma they face within their communities due to illness. The financial burden of transportation costs to clinics, along with limited access to essential resources like water and nutritious food, has made it difficult for participants to manage their health and treatment.

#### Systems-Level Challenges

32% of survey respondents reported insufficient social support in their community as a challenge related to EWEs affecting HIV/TB care (329/1024, 32%). Overall, 1071 (59%) of participants reported noticing changes in the occurrence of health issues in their communities in the past 5 years. Survey participants reported noticing increases in diarrheal infections (480, 45%), malnutrition (453, 42%), HIV and TB treatment non-adherence (479, 45%), and mental health issues (416, 39%).

Approximately half of the survey participants (1010/1804, 56%) reported being aware of any efforts in their community that helped mitigate the impact of climate disruptions on healthcare. Respondents reported being aware of food and nutrition support (679/1010, 67%), water and sanitation (543/1010, 54%), and livelihood programs (394/1010, 39%).

### Potential components for climate adaptive TB/HIV service delivery strategies

Participants discussed the need for further programs and additional support beyond what they may have required in their lives prior to these environmental changes. Participants discussed the need for community health outposts for decentralized community-based provision of health services (1086/1804, 60%), water/sanitation services (781/1804, 43%), nutrition support (541/1804, 30%), and transportation support (423/1804, 24%).

During the FGDs, areas for which participants said support would be beneficial given the added challenges of climate change and EWEs on HIV and TB care included food and water access, transportation and physical access to care support, income generating activity support, education and awareness, medication access, and climate adaptation strategies (Table 3).

**Table 3.** Strategies identified through conducting community needs assessment to address the impacts of climate change on HIV and TB care in Zimbabwe.

| Categories | Suggested interventions |
| --- | --- |
| Climate-resilient nutrition support | HIV and TB treatment being accompanied by nutrition assistance<br>Food hampers including kapenta, beans, mealie meal, tinned goods, and sugar<br>Special food assistance for those unable to work during EWEs |
| Water and sanitation | Provision of safe water sources for patients during EWEs<br>Provision of alternative safe water sources, such as drilling boreholes, establishing solar powered water systems, and installation of piped water systems<br>Maintenance of dams to increase their water-holding capacity<br>Support for maintaining health and hygiene (providing soap, clean water, a good sleeping space, and well-ventilated housing) during EWEs |
| Transportation and access | Conduct community outreach for treatment to bring healthcare services closer to communities and reducing transport costs for patients during EWEs<br>Organizing outreach programs<br>Implementing mobile clinics<br>Implement door-to-door visits for patients who are unable to travel<br>Mitigate the costs for patients by providing transportation stipends and free ambulance services<br>Improve quality of roads to ensure accessibility to health facilities during EWEs |
| Income generating activities and financial support | Support income generating projects, such as vocational training in poultry farming, gardening, baking, sewing, and goat farming to mitigate challenges due to loss of income during EWEs<br>Cash transfer programs during EWEs<br>Offer incentives for screening for TB<br>Provision of internal savings and lending schemes to mitigate the financial impacts of EWEs |
| Education and awareness | Establish community awareness programs on climate change and health |
| Medication access | Ensure a consistent and adequate supply of medication to avoid barriers to medication access during EWEs |
| Climate adaptation strategies | Provision of seeds for conservation farming activities<br>Conduct community training on conservation farming<br>Establish communal irrigation schemes |

## Discussion

Climate change increasingly undermines the social and structural determinants of health for PLHIV and those affected by TB, with direct implications for care engagement, treatment adherence, and overall wellbeing. Our formative evaluation utilized both quantitative and qualitative data to document the impacts of EWEs on both people living with HIV and those affected by TB. Our findings extend prior conceptual frameworks, including those describing climate-sensitive TB determinants^1^, climate-HIV pathways mediated through food insecurity^2^, and summarizing the influence of climate change on the risk factors for TB^4^ by providing empirical evidence from a highly climate-vulnerable setting.

The majority of TB and HIV affected individuals (76%) who participated in our survey noticed changes in the climate in their communities over the past 5 years. Of those individuals, 75% reported disruptions to their HIV care due to climate change. The convergence of food insecurity, unsafe water access, mental health strain, transportation challenges, and insufficient social support emerged as key pathways linking EWEs to HIV/TB care disruptions. These findings were consistent across FGDs, underscoring the credibility and coherence of mechanisms by which these determinants affect TB and HIV care delivery outcomes.

Food insecurity was perceived by participants to be the most influential climate-sensitive determinant of TB and HIV outcomes identified across data sources. These findings reinforce substantial prior literature characterizing food insecurity as a barrier to HIV viral suppression, a driver of opportunistic infections, and a determinant of TB progression and treatment outcomes through immune recovery, facilitated treatment adherence and overall TB treatment success.^**16-18**^ Participants described how competing demands for limited food and water resources constrained their ability to prioritize medication adherence and contributed to physical discomfort when taking treatment without food. Over 86% of participants noted that they experience food insecurity, which often disrupts their health status as adequate nutrition is essential to support immune recovery, medication adherence, and overall treatment success among individuals affected by both TB and HIV.

Climate change-related livelihood losses, water insecurity, transportation obstacles, and psychosocial stress and impoverishment further eroded individuals’ capacity to prioritize HIV/TB care through pathways that exacerbate **e**conomic vulnerabilities, reduce health system access, and precipitate psychosocial stress. These findings are also in keeping with conceptual frameworks for extreme weather events and both TB and HIV outcomes. Our analysis also underscored the interdependence between these social and environmental determinants. Conversely, the impact of EWEs on migration and displacement considered prominent factors existing frameworks^1,2^, was not a finding in our study. This may be due to contextual factors specific to our study participants in Zimbabwe, who may not be as mobile, or that we potentially undersampled individuals who may have themselves already migrated due to EWEs such as the El Niño drought.

These results highlight the need for multi-level implementation strategies that mitigate the effects of climate-related shocks on essential resources and service delivery. Suggested strategies reported by study participants included nutrition and water access support, transportation and physical access to healthcare, financial support, among other interventions. Our study participants noted that while some programs exist, such as food and nutrition support, water and sanitation, and livelihood programs, additional programs are needed to optimize inclusivity of this vulnerable population and responsiveness to their specific needs. By mapping themes from the qualitative analysis to the CFIR implementation science framework, we demonstrated that determinants which pose barriers to care at the individual, health facility and community, and system-level. Our qualitative analysis suggests that programs to mitigate the impacts of climate change on HIV and TB care could include nutrition support, water and sanitation interventions, transportation and health care access interventions, and income generating activities, among others.

One strategy frequently mentioned by participants to mitigate the effects of EWEs on access to HIV and TB care is social protection. Social protection, defined as social and economic policies targeting multiple dimensions of poverty, improves access to basic services and supports livelihoods, reducing the impact of financial shocks on vulnerable populations.^19,20^ Examples include cash, food, or transport assistance; access to social services; income-generation training; community savings groups; and human-capital development programs.^20,21^ Food or nutrition support to help mitigate the effects of malnutrition and food insecurity on people with TB is already recommended by normative guidance, although it is not consistently nor widely implemented.^22^ Such interventions can enhance nutrition and treatment outcomes among people affected by TB and PLHIV. In India, modest nutritional support improved TB treatment success^23^ while studies from South Africa and Ethiopia found that lower weight or poor nutrition increased mortality even during treatment.^24,25^ The RATIONS study in India further showed that nutritional support and weight gain reduced TB mortality risk.^26^ In the context of climate change and EWEs, food and nutritional support of longer duration and more intensity may be more urgently required for populations who experience increased vulnerability.

Limitations of this research include a limited number of FDGs across different subgroups (HIV and TB affected individuals) which may have limited data saturation, the absence of full transcription due to resource constraints, and that the study was conducted in a single country, Zimbabwe, a context that may not fully represent climate vulnerability patterns elsewhere. However, Zimbabwe exemplifies the intersection of high TB/HIV burden and climate fragility, positioning it as an important sentinel setting for understanding climate-health interactions. In addition, this study has several strengths, including the use of both quantitative and qualitative methods which allowed us to validate results and provide richer context, triangulation across multiple impacted populations including PLHIV, people affected by TB, and the healthcare providers who care for them; and use of implementation science frameworks to identify mechanisms.

Future research should build on these findings by quantifying the magnitude and mechanisms through which climate change affects TB and HIV outcomes. Specifically, studies are needed to disentangle the relative impacts of distinct extreme weather events, such as the 2023/2024 El Niño-induced drought, on service delivery, treatment adherence, and health outcomes. Implementation science approaches will be critical to develop and test adaptation strategies that strengthen health system resilience and continuity of care under climate stress. This includes identifying which social protection, nutrition, and livelihood interventions are most effective and sustainable in high-burden, resource-constrained settings. Finally, expanding this work beyond Zimbabwe to other regions experiencing overlapping TB, HIV, and climate vulnerabilities will be essential to ensure generalizability and to guide evidence-based policy and programmatic responses.

## Conclusion

Extreme weather events caused by climate change create a cascade of challenges that hinder individuals’ ability to adhere to HIV and TB treatment. In Zimbabwe, extreme heat and drought are impacting food and water security as well as financial implications creating challenges for patients to access care. Findings underscore the urgent need for integrated, climate-adaptive social protection strategies to maintain HIV and TB care continuity in climate-vulnerable regions like Zimbabwe.

## Data Availability

All data produced in the present study are available upon reasonable request to the authors.

## Notes

### Competing Interest Statement

The authors have declared no competing interest.

### Funding Statement

This study was funded by the Nina Ireland Program for Lung Health (PI: Shete) and the National Center for Advancing Translational Sciences, National Institutes of Health, through UCSF-CTSI Grant Number UL1 TR001872.

### Author Declarations

The Institutional Review Boards of the Medical Research Council of Zimbabwe and the University of California, San Francisco gave ethical approval for this work.

