## Appendix A for "Assessing the Impact of Extreme Weather Events on HIV and Tuberculosis Care in Zimbabwe"

| Table 1. Focus Group Discussion Guide for Recipients of TB Care | | |
| --- | --- | --- |
| **TOOL 1** | **Narrative invitation** | **Further questions** |
|  | **Thank you very much for taking your time to participate in this group discussion. With this group discussion, we aim to get a clearer picture about important information on the impacts of climate change on health, particularly TB care and treatment, and would help identify the most pressing needs for the community. You may all have different views about this, but I’d like to remind you that there are no right or wrong answers and that I’d like to hear all of your opinions, so feel free to each tell me what you think.** | |
| 1.1 | **Can you share your experiences or thoughts on how climate change, such as drought, flooding, or increased temperatures, has impacted your health and overall wellbeing?** | - *Have you noticed any changes in your ability to access TB treatment, healthcare services, or medications due to extreme weather events or changing climate patterns?* - *What challenges have you faced in following your TB treatment regimen (e.g., taking medication regularly, completing the full course of treatment) because of climate change like food shortages, transportation issues, or access to healthcare facilities?* |
| 1.2 | **What are the key barriers related to climate change that has prevented you from consistently accessing healthcare for TB?** | - *Have you experienced any difficulties accessing TB medications, either due to supply issues or transportation problems related to climate disruptions? If yes, how did this affect your treatment?* |
| 1.3 | **How has climate change made it difficult for you to afford TB treatment or the costs associated with healthcare (e.g., transport to the clinic, user fees)?** | - *Have you had to make difficult choices between spending money on your TB treatment or meeting other basic needs (such as food, water, rentals) because of climate-related challenges?* - *What are some of the social impacts of climate change that you feel are affecting your ability to manage your TB treatment and overall health?* |
| 1.4 | **How has the changing climate (e.g., drought, erratic rainfall) affected the availability of food in your community? How has this impacted your health, especially your ability to maintain a healthy diet during TB treatment?**  **What support do you think would be most helpful to ensure that TB patients in your community have consistent access to food and nutrition, especially during climate change?** | - Have there been any instances where you were unable to access adequate nutrition while on TB treatment due to climate-related challenges (e.g., food shortages, rising food prices, or poor harvests)? |
| 1.5 | **What strategies or practices has your community adopted to cope with the effects of climate change, especially in relation to health and TB care?** | - *What do you think your community can better adapt to the changing climate in order to ensure better healthcare access for TB patients?* - *What support from the government or health organizations do you think would be most effective in helping TB patients navigate the challenges posed by climate change?* |
| 2.1 | **Do you think there are specific water and sanitation challenges that need to be addressed to help prevent the spread of TB in your community, especially in the context of climate change?** | - *How has this affected your ability to manage TB, especially in terms of hygiene and infection control?* |
| 2.2 | **What specific government or community-based interventions would help improve the management of TB in your community in the face of climate change?** | - *In your opinion, how can health systems in Zimbabwe better prepare for the intersection of climate change and TB care?* - *What role do you think local health workers or community leaders can play in mitigating the impacts of climate change on TB care and treatment?* |
| 2.3 | **How can TB patients and communities in Zimbabwe better prepare for future climate change?** | - *What long-term changes would you like to see in your community to better address the health impacts of climate change, particularly for people living with TB?* |
|  | **Given our focus, do you have additional information, ideas or suggestions that were not discussed during this discussion that you would like to share with us?**  **Thank you all for this discussion!** | |

| **Table 2.** Focus Group Discussion Guide for Recipients of HIV Care | | |
| --- | --- | --- |
| **TOOL 1** | **Narrative invitation** | **Further questions** |
|  | **Thank you very much for taking your time to participate in this group discussion. With this group discussion, we aim to get a clearer picture about important information on the impacts of climate disruptions on health, particularly HIV care and treatment, and would help identify the most pressing needs for the community. You may all have different views about this, but I’d like to remind you that there are no right or wrong answers and that I’d like to hear all of your opinions, so feel free to each tell me what you think.** | |
| 1.1 | **Can you share any changes you've noticed in the weather in your community? How have these changes impacted your daily life or your community.?** | - *How do you think these changes in weather have affected your health and the way you manage your HIV care?* - *Can you share how climate change (e.g., heatwaves, droughts, flooding) has affected the availability of food, water, and other essential resources in your community?* |
| 1.2 | **What challenges have you encountered in accessing HIV care or treatment during extreme weather events, such as heatwaves, floods, or droughts?** | - *Has the availability of your HIV medications been affected during climate-related events? If yes, how?*   *(For example: medication stock-outs, inability to access healthcare facilities, difficulty storing medication due to high temperatures, etc.)*   - *How has transportation to healthcare facilities been affected by adverse weather conditions?* - *Have you had trouble keeping up with your regular HIV care appointments because of these climate-related challenges?* |
| 1.3 | **Has your financial situation been affected by climate change, particularly in terms of affording HIV care or related services (e.g., transportation, medication)?** | - *Have you ever had to miss a healthcare appointment or go without treatment because you couldn’t afford the associated costs due to climate disruptions?* - *How do climate-related events (like droughts or floods) impact your ability to buy food, access healthcare, or manage transportation costs?* - *What types of support (financial or material) would help you continue to access HIV care during climate-related events?* |
| 1.4 | **How important is nutrition for your HIV care, and have you experienced challenges in accessing nutritious food during climate change?** | - *Has climate change impacted the availability of food or water in your community? How has that affected your ability to stay healthy and continue your HIV treatment?* - *Do you receive any support (e.g., food assistance, nutrition programs) to help you manage nutrition during times of climate stress? If yes, was it helpful?* |
| 1.5 | **What strategies have you used to cope with climate-related challenges to your HIV care (e.g., finding alternative healthcare services, using public transportation, adjusting your medication schedule, etc.)?** | - *How do you manage disruptions to your HIV treatment or medication supply during difficult times (e.g., climate-related shortages, infrastructure damage)?* - *Have you or others living with HIV developed specific ways of dealing with the challenges posed by climate disruptions, such as seeking help from others or accessing community-based resources?* |
| 2.1 | **What kind of support would be most helpful to you in managing your HIV care during periods of climate change? (For example: transportation assistance, increased multi-month dispensing of medication, financial aid, food support, etc.)** | - *Have you benefited from any community-based programs that assist people living with HIV during climate-related challenges? If yes, please describe how they helped.* - *Do you think providing cash transfers, livelihood projects, or food assistance would reduce the impact of climate disruptions on your health? Why or why not?* |
| 2.2 | **Do you think your community is aware of the impact of climate change on health, especially in relation to HIV?** | - Have you had any discussions or educational sessions about climate change and its effect on HIV care? If not, what kind of information would be helpful for you? - What can health services or local authorities do to better educate people living with HIV on the impacts of climate change and how to adapt? |
|  | **Given our focus, do you have additional information, ideas or suggestions that were not discussed during this discussion that you would like to share with us?**  **Thank you all for this discussion!** | |
